# COVID-19 Asymptomatic Infection Estimation

**DOI:** 10.1101/2020.04.19.20068072

**Authors:** Yang Yu, Yu-Ren Liu, Fan-Ming Luo, Wei-Wei Tu, De-Chuan Zhan, Guo Yu, Zhi-Hua Zhou

## Abstract

**Background:** Mounting evidence suggests that there is an undetected pool of COVID-19 asymptomatic but infectious cases. Estimating the number of asymptomatic infections has been crucial to understand the virus and contain its spread, which is, however, hard to be accurately counted.

**Methods:** We propose an approach of machine learning based fine-grained simulator (ML-Sim), which integrates multiple practical factors including disease progress in the incubation period, cross-region population movement, undetected asymptomatic patients, and prevention and containment strength. The interactions among these factors are modeled by virtual transmission dynamics with several undetermined parameters, which are determined from epidemic data by machine learning techniques. When MLSim learns to match the real data closely, it also models the number of asymptomatic patients. MLSim is learned from the open Chinese global epidemic data.

**Findings:** MLSim showed better forecast accuracy than the SEIR and LSTM-based prediction models. The MLSim learned from the data of China’s mainland reveals that there could have been 150,408 (142,178-157,417) asymptomatic and had self-healed patients, which is 65% (64% – 65%) of the inferred total infections including undetected ones. The numbers of asymptomatic but infectious patients on April 15, 2020, were inferred as, Italy: 41,387 (29,037 – 57,151), Germany: 21,118 (11,484 – 41,646), USA: 354,657 (277,641 – 495,128), France: 40,379 (10,807 – 186,878), and UK: 144,424 (127,215 – 171,930). To control the virus transmission, the containment measures taken by the government were crucial. The learned MLSim also reveals that if the date of containment measures in China’s mainland was postponed for 1, 3, 5, and 7 days later than Jan. 23, there would be 109,039 (129%), 183,930 (218%), 313,342 (371%), 537,555 (637%) confirmed cases on June 12.

**Conclusions:** Machine learning based fine-grained simulators can better model the complex real-world disease transmission process, and thus can help decision-making of balanced containment measures. The simulator also revealed the potential great number of undetected asymptomatic infections, which poses a great risk to the virus containment.

**Funding:** National Natural Science Foundation of China.

## 1 Introduction

The virus, named as COVID-19, which was identified in Wuhan city in December 2019, is a coronavirus and belongs to the same family as the pathogen that causes severe acute respiratory syndrome (ARDS), or SARS [Li *et al*., 2020; Zhou et al., 2020]. It causes a respiratory dominated illness and can spread from person to person [Chan *et al*., 2020; Wang et al., 2020b; Xu et al., 2020].

Asymptomatic cases refer to people who can be tested positive for the coronavirus but develop limited or no symptoms such as fever, cough, or sore throat, noting that they are infectious and pose a risk of spreading to others. Mounting evidence suggests that there is an undetected pool of covert asymptomatic cases [Dong *et al*., 2020; Wang et al., 2020a]. A recent news [Qiu] published by Nature on March 20, 2020, manifests that there could be 30% – 60% patients are asymptomatic or mildly ill cases. Estimating the number of undetected asymptomatic cases has been crucial to containing the spread of the coronavirus, which is, however, hard to be accurately counted. Meanwhile, if we can model how the virus transmits, it is fully possible to make inference on the unobserved number of asymptomatic patients from the observed epidemic data.

There are mainly two ways to model the virus transmission. One way is to employ transmission dynamics to describe how diseases spread. A classical transmission dynamics model is the Susceptible Infected Recovered (SIR) model [Kermack and McKendrick, 1991] that consists of susceptibles, infectives, and recovered individuals, and differential equations about how individuals changes. Variants of SIR models have been studied, such as the SEIR model [Anderson *et al*., 1992; Lekone and Finkenstädt, 2006] considering the incubation period, which is also significant for COVID-19 infection, and other models considering temporary immunity [Wen and Yang, 2008], passive immunity [Bichara *et al*., 2014; Qureshi and Yusuf, 2019], etc. These models are grounded in human knowledge about virus infection and immunity, which are good in generalization to long-period predictions. At the same time, these models commonly over-simply the real-world and are hard to fit well the epidemic data, which results in large prediction errors. Another way is to rely on machine learning models. For epidemic data, a type of model that are naturally suitable for the task is the recurrent neural networks (RNNs) [Elman, 1990], with long short-term memory (LSTM) [Hochreiter and Schmidhuber, 1997]. These models are very flexible that can fit well the epidemic data, and thus can make accurate predictions for the very near future. However, due to the lack of domain knowledge, these models are hard to generalize to long-term futures, hard to incorporate different decisions, and hard to be interpretable.

Noticed that combination of human knowledge and learning from data has recently shown powerful in solving sophisticated problems [Zhou, 2019]. To alleviate the above issues of the two types of methods, we propose an approach of machine learning based fine-grained simulator (MLSim), which can not only predict the virus transmission more accurately, but also help to estimate the number of asymptomatic patients.

## 2 Method

We propose a machine learning based transmission simulator (MLSim) to estimate COVID-19 asymptomatic infections. MLSim is data-driven and integrates multiple crucial factors, including disease progress in the incubation period, cross-region population movement, undetected asymptomatic patient numbers, and prevention and containment strength. The source code of MLSim is publicly accessible at https://github.com/eyounx/MLSim.

### 2.1 Simulator structure

The simulator structure, which is designed according to the transmission characteristics of COVID-19, is shown in Figure 1. The disease progress is divided into three stages: incubation stage, quarantine stage, and confirmation stage. All three stages are represented as fixed-length lists, whose length is determined by the clinical experience. Each number in the list indicates the number of patients on that day. The patients in these stages are respectively called latent patients, quarantined patients, and confirmed patients. Latent patients are infectious and temporarily mildly ill or asymptomatic. Once they show obvious symptoms, they will be quarantined and lose the infectivity.

**Figure 1:**
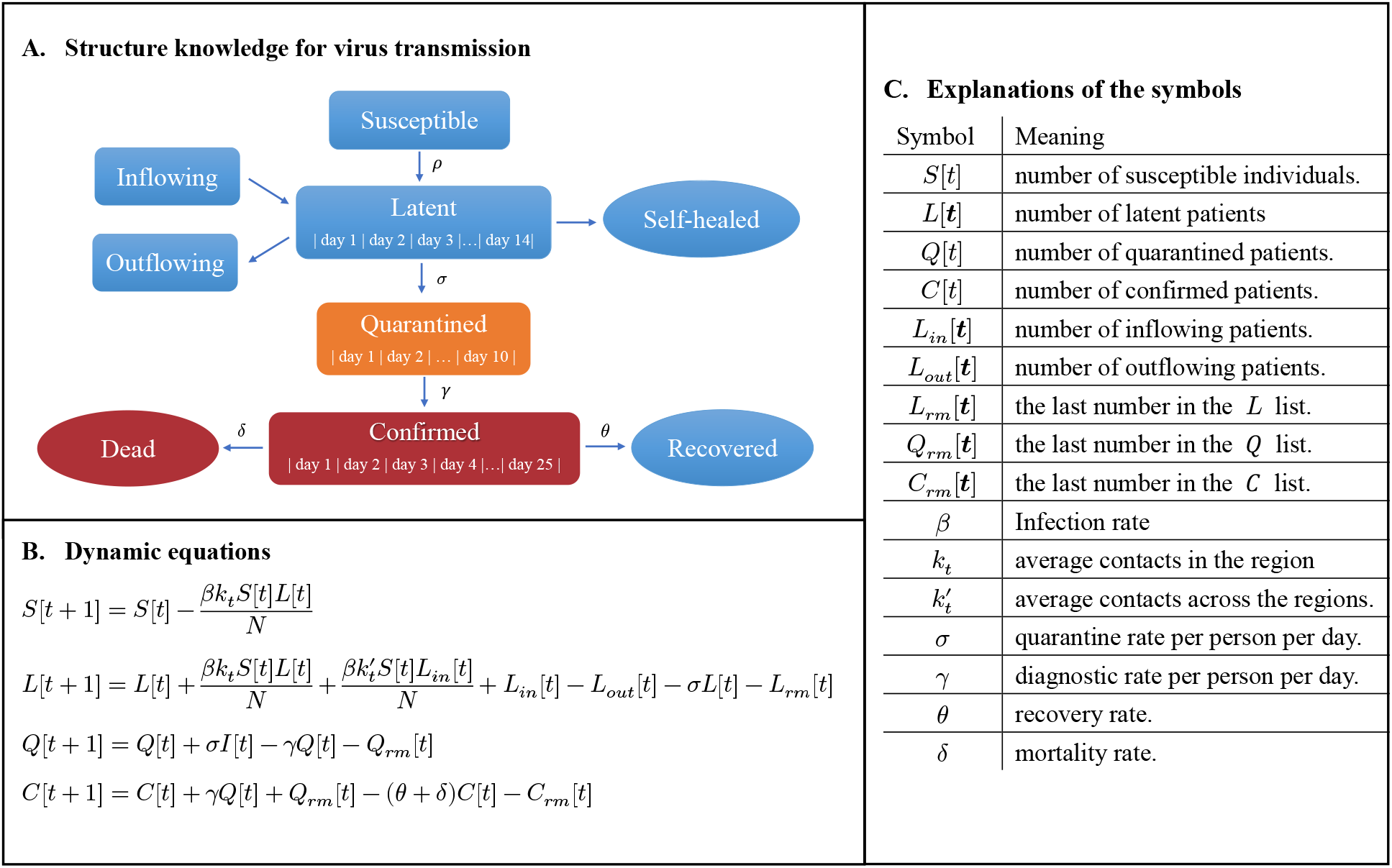
Model the virus transmission. MLSim is built based on the transmission characteristics of COVID-19. The infections in MLSim are divided into latent, quarantined, and confirmed patients. Only latent patients have the ability to infect others. Each kind of patients can experience a period of time to move to the next stage. Latent patients can show obvious symptoms and be quarantined with the probability of *σ* on any day of 14 days, which is the time of the incubation period. The latent patients who have experienced the whole incubation period and are not quarantined are usually mildly ill or asymptomatic. We assume these patients will self-heal. The self-healed patients were not recorded in the published data, and calculating their number can help to estimate the number of asymptomatic patients. See Appendix A for more explanations of why self-healed patients are essential. The parameters in MLSim are learned through derivative-free optimization since the simulator is non-differential.

In each day, the susceptible populations can be infected and turned into latent patients. Meanwhile, the external latent patients can infect other passengers through the cross-region population movement. The number of newly infected patients will be appended as the first day in the latent patient list, and thus the number of patients each day in the list automatically moves to the next day. Then, the number on the last day of the list will be removed, which means the patients that the number represents have experienced the whole incubation stage, i.e., 14 days, without showing obvious symptoms and being quarantined. They are assumed to be self-healed. The latent patients can show obvious symptoms and be quarantined on any day of the incubation stage. The quarantined patients will be confirmed according to the diagnostic rate on any day of the quarantine stage or when they are moved out of the list. It should be noticed that latent patients may not be quarantined, but all quarantined patients will be confirmed. The mortality/recovery rate represents the probability of death/recovery per day. If the confirmed patients have experienced the whole confirmation stage, i.e., 25 days, they are assumed to have recovered.

### 2.2 Determining simulator parameters

To finalize the simulator, we need to determine the parameters. Our goal is to find the parameters that make the simulation outcome as similar as possible with the real epidemic data. In the following parts, we introduce the parameters to be optimized, the loss function, and the optimization process.

#### 2.2.1 Parameters

We initialize the simulator by setting *S*[0] = *N* (*N* is the population of the region), *Q*[0] = 0 and *C*[0] = 0. We have 8 parameters to be optimized, respectively are *β, k k*′, *σ, γ, θ, δ* and *I*(0). The start date of the simulator was 14 days before the date when the epidemic data was first released. Because the real number of latent patients on that day is unavailable, we leave it to be optimized (*I*[0]). We assume the average number of individual contacts per person per day before containment is 15. *k* represents the average number of individual contacts within a region after the containment. *k*′ represents the average number of individual contacts in the cross-region population movement after the containment. 15/*k* and 15/*k*′ reflect the government’s containment strength. A smaller *k* indicates a higher containment strength. The explanation of other parameters is the same as that of Figure 1.C.

#### 2.2.2 Loss function

Let the difference between the real number of newly confirmed cases on the *t*^th^ day and its simulated counterpart be Δ*N*_*c*_[*t*]. Analogously, let the difference between the real number of new deaths and its simulated counterpart on the *t*^th^ day be Δ*N*_*d*_[*t*] and the difference between the real number of new recoveries and its simulated counterpart on the *t*^th^ day be Δ*N*_*r*_[*t*]. Then the loss function, which maximizes the utilization of all public data, is defined as:

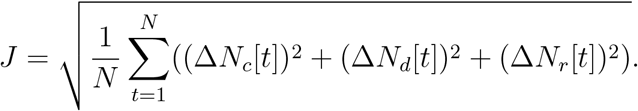

#### 2.2.3 Optimization

Since the rules of the simulator are non-differentiable, the parameters can not be optimized through derivative-based methods, e.g., stochastic gradient descent.

Derivative-free optimization, also termed as zeroth-order or black-box optimization, involves a class of optimization algorithms that do not rely on gradient information. Typical derivative-free optimization algorithms include evolutionary algorithms [Hansen *et al*., 2003; Larrañaga and Lozano, 2001; Neumann and Wegener, 2007], Bayesian optimization [Kawaguchi *et al*., 2015; Martinez-Cantin, 2014; Wang et al., 2016] and recently emerged classification-based optimization methods [Hu *et al*., 2017; Liu et al., 2019; Yu et al., 2016]. ZOOpt [Liu *et al*., 2018] is a python package for derivative-free optimization. It implements some state-of-the-art classification-based derivative-free optimization methods and their parallel versions, which can quickly approximate the optimal solution to the problem. We used ZOOpt to obtain the parameters that minimize the loss function.

## 3 Experiments

This section evaluates the validation of the MLSim approach.

### 3.1 Data sources

The most recent epidemic data based on daily COVID-19 outbreak numbers were retrieved from two open-source GitHub repositories, which respectively share the Chinese epidemic data ^1^ and the global epidemic data ^2^. The abrupt increase of confirmed cases in China on Feb. 13 is exponentially averaged onto the numbers in the previous 32 days. The population movement data in China’s mainland were sourced from baiduqianxi^3^, which gives the migration index based on the daily number of inbound and outbound events by rail, air, and road traffic.

### 3.2 Evaluations

We first tested the prediction accuracy of MLSim and compared it with the SEIR and LSTM-based model.

The COVID-19 data of China was divided into two parts. The first part, the training data, included the data before Feb. 10, 2020. And the second part, the validation data, included the rest. The start date of the simulator was set to be Dec. 28, 2019. The number of inflowing (outflowing) patients can be calculated as 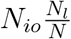, where *N*_*io*_ is the inflowing (outflowing) population size, *N*_*l*_ is the number of latent patients in the source region and *N* is the population of the source region.

We constructed a simulator for each China province. The simulator parameters of Hubei were first optimized, where the inflowing patients from the remaining provinces were not considered. Because *k*′·*I*_*in*_[*t*] was always equal to 0, the parameter *k*, average individual contacts in the cross-region population movement, was not optimized. Then we fixed the parameters of Hubei and separately optimized the parameters of the remaining China provinces, which only considered the inflowing patients from Hubei. The parameters of the SEIR and LSTM model were obtained by fitting the training data, i.e., minimizing the mean square error between the predicted results and the real data. Details of the training can be found in Appendix B and Appendix C.

Models were also trained on the data of South Korea. The training data included the data before Mar. 5, and the validation data included the rest. The COVID-19 data of China was added for training the LSTM model.

The results are shown in Figure 2. It can be observed that MLSim achieved the best validation performance in all cases. Notice that the training data of Hubei has not shown an evident decline yet. The LSTM model gave a constant prediction. In Figure 2-D, the curve generated by the LSTM model shows a periodic characteristic. The curve generated by the LSTM model in Figure 2-F is similar to the training data of China, which shows that a small increase of training data cannot help LSTM learn something essential but only makes it remember more data. The generalization performance of the SEIR model is worse than that of MLSim in all cases.

**Figure 2:**
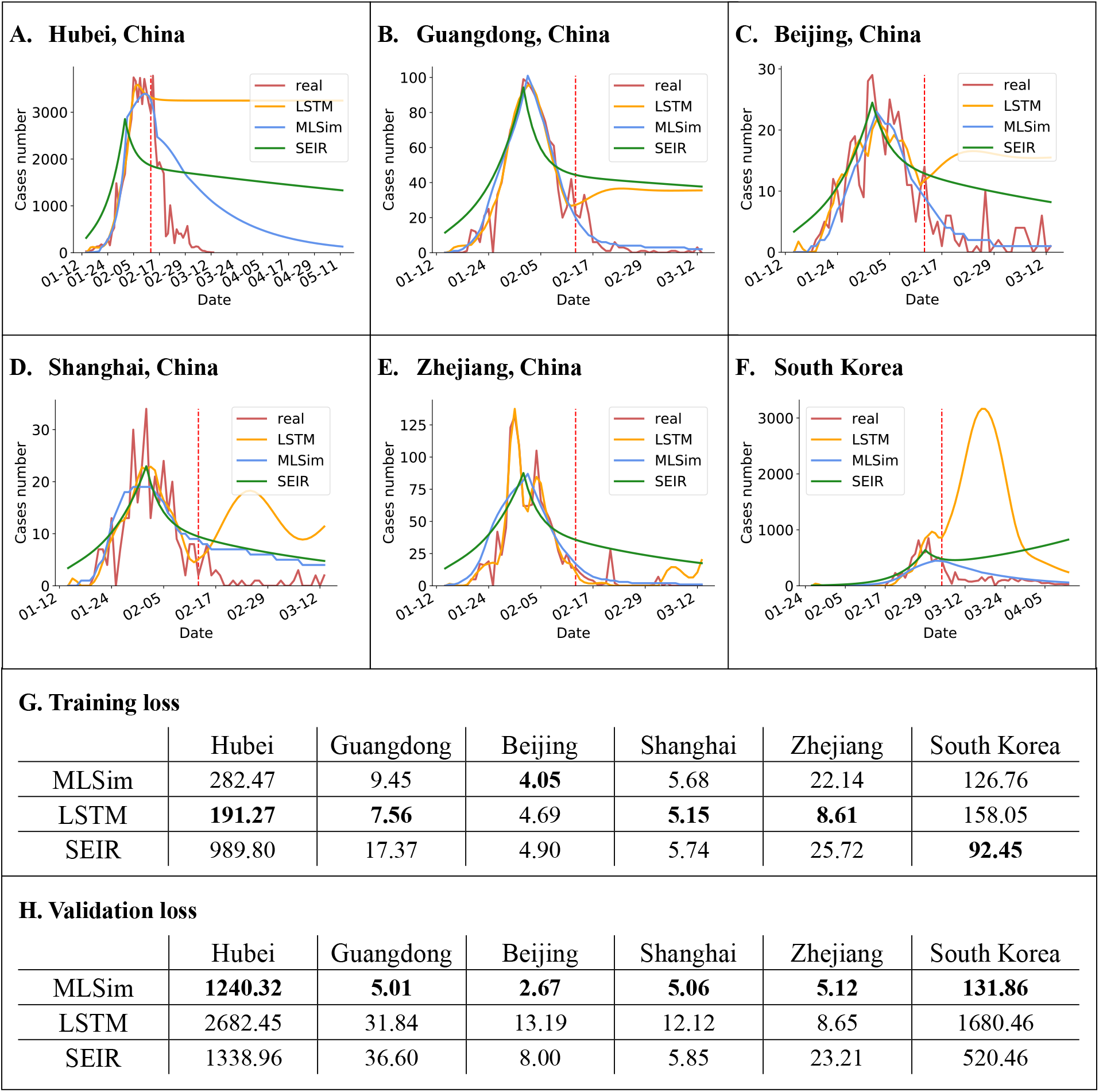
Evaluate the forecast accuracy. MLSim was compared with the LSTM model and the SEIR model. The red vertical dash line denotes the date when the data was split into training and validation data.

The above results demonstrate that MLSim can forecast virus transmission more accurately. The SEIR model is much simpler, which leads to a comparatively weak representation ability. Neural networks like LSTM have strong representation ability, but it can only predict the near future for the lack of domain knowledge and suffers from the overfitting problems when the amount of available data is limited.

## 4 Interpretations

### 4.1 Learning results

We obtained the simulator parameters of 31 China provinces and 6 other countries by fitting the data. The optimization was repeated 10 times, and for each parameter, the median value and its 95% confidence interval are recorded. Here we discuss the results of Hubei. The results of other provinces and countries can be found in Appendix F. For Hubei, the infection rate *β*, i.e. the rate of transmission for the susceptible to be infected, is 0.023 (0.018-0.027) and the quarantine rate *σ*,; i.e. the rate by which the latent patients develops obvious symptoms and are quarantined per day, is 0.030 (0.030-0.031). The initial number of infected patients on Dec. 28, 2019 is 106 (29-397). The average individual contacts has decreased by 87% (83%-90%) (calculated by 1−*k*/15) since Jan. 23, 2020, the start date of containment in China. The reproductive number, which means the number of cases one case generates on average over the course of its infectious period, can be calculated as 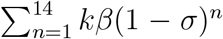, where *k* is the average number of individual contacts, 14 is the maximum length of incubation period set in our simulator. The reproductive number before and after the containment are respectively equal to 3.850 (3.083-4.595) (*R*_0_) and 0.499 (0.489-0.520) 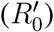. Ignoring the self-healed patients, the epidemic doubling time in the early stage can be calculated as 1/ log2(1 + *kβ*(1 −*σ*)), which is equal to 2.625 (2.217-3.267) days. The obtained doubling time is shorter than that estimated in some previous work [Kucharski *et al*., 2020; Wu et *al*., 2020a,b] because we considered the undetected patients in this paper.

### 4.2 Estimate the number of undetected asymptomatic cases

MLSim assumes that latent infections are currently asymptomatic and if they don’t show obvious symptoms and be quarantined in the whole incubation period, they will self-heal. Figure 3 shows the simulated results of MLSim. We found that only 35% (35%-36%) infections were detected in China and 65% (64%-65%) infections were asymptomatic and had self-healed. The current numbers of latent patients in South Korea, Italy, Germany, USA and UK respectively are 112 (40-262), 41,387 (29,037-57,151), 21,118 (11,484-41,646), 354,657 (277,641-495,128), 144,424 (127,215-171,930), posing a great risk to the virus containment.

**Figure 3:**
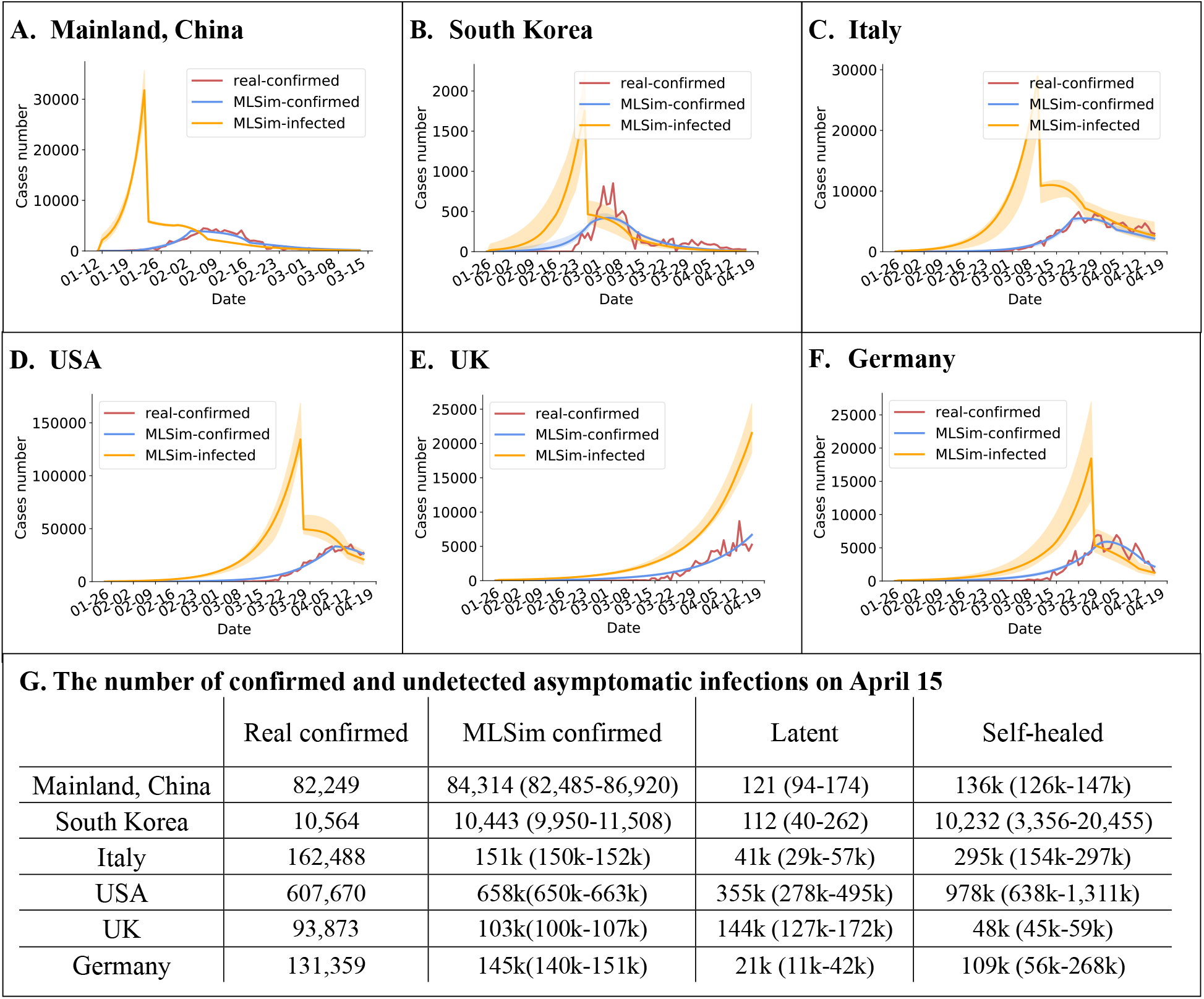
Estimate the number of asymptomatic cases. MLSim assumes that latent infections are currently mildly ill or asymptomatic. If they don’t develop obvious symptoms and be quarantined in the whole incubation period, they will self-heal. The number of total infections is the sum of the number of confirmed, quarantined, latent and self-healed cases. The abrupt decrease of the number of new infections in countries except UK is due to the learned average individual contacts after containment (*k*). For UK, *k* was not optimized because the data were not sufficient enough to estimate the influence of containment accurately. Panel G shows the number of different kinds of infections on April 15.

### 4.3 Estimate the influence of containment

The Chinese government has carried out strict prevention and containment measures since Jan. 23, 2020 [Cao and Zhou; Paul Mozur and Krolik], which have effectively controlled the virus spread but caused inevitable economical loss [The Economist]. What if the measures were not taken? We use the optimized simulator to find the answer. We simulated the virus transmission retrospectively with different containment start dates and containment-strengths.

We first postponed the start date of containment. The simulation results are shown in Figure 4, which demonstrate that there would be 109,039 (129%), 183,930 (218%), 313,342 (371%), 537,555 (637%) confirmed cases on June 12 if the start date of containment was postponed by 1, 3, 5 or 7 days.

**Figure 4:**
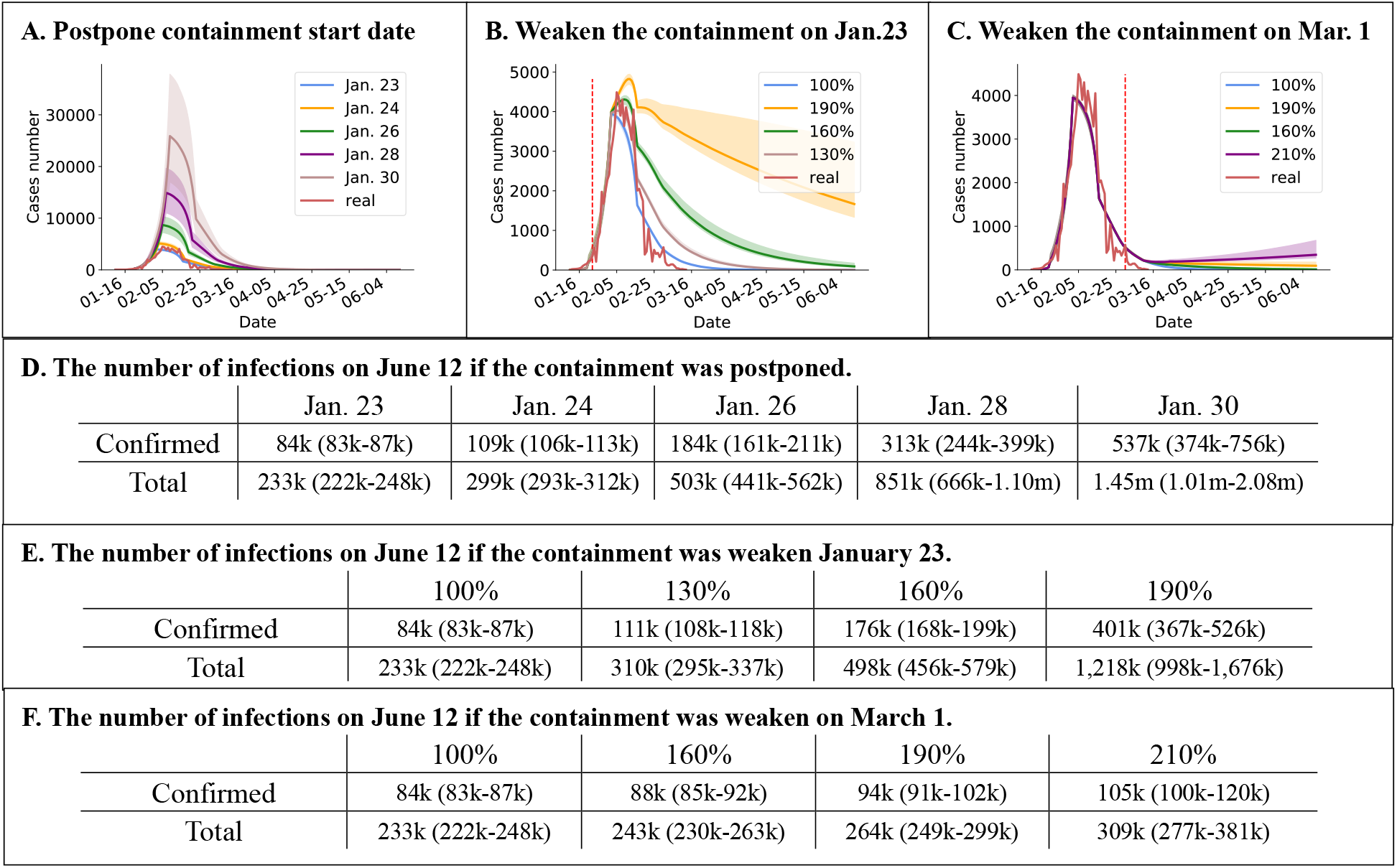
What if the containment was postponed or weakened in China? MLSim can be useful to evaluate and facilitate containment decisions, by answering “what-if” questions. The red vertical lines in panel B and C denote the date when the containment strength was changed. Legends in panel A denote the start date of the containment. If the containment was weakened, the average individual contacts after the containment (*k*) will increase. Legends in panel B and C and percentages in the first line of table E and F denote how much *k* was increased to if the containment was weakened on Jan. 23 and Mar. 1.

The prevention and containment measures will directly change the value of *k*, i.e. the average number of individual contacts. A greater containment-strength causes a smaller *k*. Therefore, the containment-strength can be relaxed by increasing *k* value while leaving other parameters unchanged. We increased *k* by 30%, 60%, 90% on Jan. 23 and by 60%, 90%, 110% on March 1 respectively to investigate their influence. It can be observed that if *k* was increased by 30% on Jan. 23, the cumulative number of confirmed cases would increased to 111,460 (132%) on June 12. If *k* was increased by 60% on Jan. 23, the cumulative number of confirmed cases would increase to 176,053 (209%). Compared with that, the influence of relaxing the containment after March 1 is much less. An increase of *k* by 60% causes 3,296 (4%) more confirmed cases. While an increase of *k* by 110% will cause a second outbreak.

Through this retrospective simulation, we conclude that the strict containment measures taken after Jan. 23 are crucial for suppressing the virus spread and a postponement of few days or slight relaxation of containment strength on that day may cause much more confirmed cases. Comparatively, an appropriate relaxation (60%) of the containment on March 1 can benefit the national economy with few adverse effects. But a great relaxation (110%) on March 1 can still cause a second outbreak.

### 4.4 Forecast the global virus transmission

The COVID-19 infections data indicates the epidemic in China’s mainland is close to an end. However, the trend in global is currently climbing. Here, we forecast the virus transmission in some countries (the obtained simulator parameters are shown in Appendix F). Inflowing patients were not considered in this experiment and its effect was transformed to the number of initial infections on Jan. 9, 2020, due to the lack of the global population movement data.

For countries except UK, the average individual contacts after the containment (*k*) can be learned by fitting the data. We forecast the virus transmission in these countries by keeping simulator parameters unchanged. While for UK, *k* cannot be learned accurately because the current data is not sufficient enough. Instead, we assumed *k* = 15 all along and forecast the future by decreasing *k* to different values. Results are shown in Figure 5. There could be 10,536 (10,051 – 11,601), 195,000 (183,770 – 247,865), 166,258 (163,497 – 171,316), 1,033,717 (994,147 – 1,120,326) and 478,117 (323,082 – 581,761) confirmed cases in South Korea, Italy, Germany, USA and France on July 15. For UK, if the containment measures were not taken, there could be 30 millions (25m-34m) confirmed cases on July 15, which means 44% (37% – 50%) populations will get infected.

**Figure 5:**
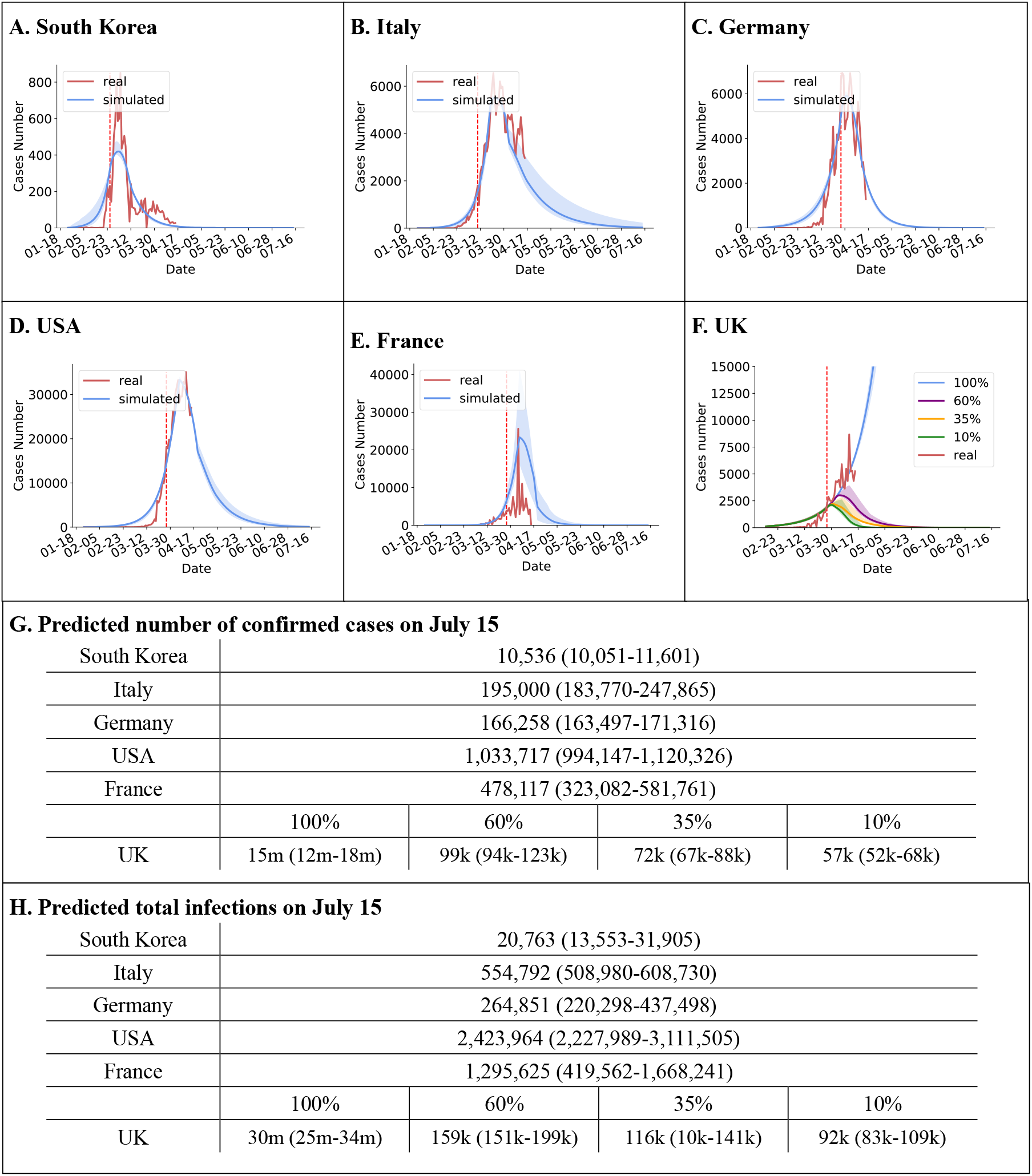
Forecast the global virus transmission. The red vertical lines denote the start date of containment. For South Korea, Italy, Germany, USA and France, the average individual contacts (*k*) after the containment was learned by fitting the data. While for UK, *k* cannot be learned accurately because the data was not sufficient enough. Instead, we assumed *k* = 15 all along and forecast the future with different levels of containment. The legends in panel F indicate the value to which *k* was decreased. For example, 35% means *k* was decreased to 35% after the containment. For reference, after containment, *k* was decreased to 13% (10% – 17%) in Hubei, China. Panel G and H demonstrate the number of confirmed cases and total infections on July 15.

## 5 Discussion

Traditional virus transmission models usually make more assumptions than MLSim and left only few parameters to be determined. For example, SEIR model had to assume the number of initial patients, incubation period and the change of average contacts after the containment in advance to determine the infection rate or *R*_0_. Although these assumptions are mostly based on clinical experience, fixing these parameters to a certain value can still be very subjective. Instead of fixing these parameters, MLSim only assumed the maximum length of each disease stage and moderately limited the search space of the parameters, rather than fixing these parameters to a certain value. Having more parameters to be optimized enables MLSim a better representation ability than traditional models. However, it also causes a side effect: there could be a multi-solution problem, in another word, there could be more than one set of simulator parameters that can fit the data and some parameters might be unreasonable. There is actually a trade-off here: the more strict the assumptions, e.g. fixing parameters to one value, the less likely the multi-solution problems could happen and the poorer representation ability the model has. An appropriate limitation on the search space of parameters can benefit the model with sufficient representation ability and fewer unreasonable solutions. Appendix D shows the settings of the search space of parameters in all experiments. In general, the multi-solution problem exists but can be eliminated by introducing more domain knowledge, i.e. more strict assumptions.

There are other potentially important factors we did not consider in our simulator, mainly due to the lack of data. The factors can include the capacity of daily tests of infections, which upper bounds the daily maximum confirmation number, and the ICU capacity, which directly affects the death rate.

## 6 Conclusion

In this paper, we proposed a machine learning based fine-grained simulator (MLSim), which built a simulator from expert domain knowledge together with learning from data. We applied MLSim to COVID-19 data and the obtained parameters can reflect its transmission characteristics. The empirical studies showed that MLSim not only can have a better long-term prediction accuracy, but can also help to estimate the number of asymptomatic infections. This kind of hybrid knowledge and data learning approach was not widely recognized in machine learning community. But we found it very useful when the data is scarce while knowledge is rich but inaccurate, such as the situation of a new contagion outbreak.

## Data Availability

all data used in this paper is publically available.

## 7 Acknowledge

We thank Mr. Su Lu, Mr. Xin-Chun Li and Mr. Hong-Wei Wei for providing baseline implementations, and Mr. Chao Wang and Mr. Yu-Yang Huang for running a daily updating system of the proposed approach for a long-term validation.

## A The impact of asymptomatic patients to the model

In this section, we try to find out if the setting of undetected asymptomatic patients is necessary. MLSim can estimate the ratio of undetected asymptomatic patients by computing (1 −*σ*)^14^, i.e., the number of self-healed patients, where *σ* is the quarantine rate. Theoretically, if the learned *σ* is large, the asymptomatic ratio will be low. For example, when *σ* is respectively equal to 0.1, 0.2 and 0.3, the asymptomatic ratio will be 23%, 4% and 0%. We limited the lower bound of the search space of *σ* to be 0.1, 0.2, 0.3 to see whether MLSim can still well fit the published data. The results are shown in Table 1. Note that the obtained parameters for Hubei (0.1, 0.2, 0.3) are inaccurate because the quarantine rate is overestimated. It can be observed that the RMSE values of these optimizations are worse than those without lower bound limitations. The results indicate that undetected asymptomatic patients are necessary to fit the real data.

**Table 1:** The obtained parameters when limiting the number of asymptomatic patients. We limited the lower bound of the search space of *σ*, the quarantine rate, to be 0.1, 0.2, 0.3 to see whether MLSim can still well fit the published data. When *σ* is respectively equal to 0.1, 0.2 and 0.3, the asymptomatic ratio will be at most 23%, 4% and 0%. Note that the obtained parameters are inaccurate for Hubei (0.1, 0.2, 0.3) because the quarantine rate is overestimated.

| | $\beta$ | $k$ | $\gamma$ | $\delta$ |
| --- | --- | --- | --- | --- |
| Hubei | 0.023 (0.018-0.027) | 1.950 (1.572-2.533) | 0.000 (0.000-0.000) | 0.005 (0.005-0.005) |
| Hubei (0.1) | 0.021 (0.021-0.029) | 3.988 (2.727-4.085) | 0.000 (0.000-0.000) | 0.004 (0.004-0.004) |
| Hubei (0.2) | 0.025 (0.024-0.025) | 6.503 (6.432-6.740) | 0.000 (0.000-0.000) | 0.004 (0.004-0.004) |
| Hubei (0.3) | 0.029 (0.029-0.034) | 8.871 (7.706-8.990) | 0.000 (0.000-0.000) | 0.003 (0.003-0.004) |
| | $I(0)$ | $\sigma$ | $k'$ | $\theta$ |
| Hubei | 106 (29-397) | 0.030 (0.030-0.031) | 4.246 (1.430-12.966) | 0.049 (0.042-0.054) |
| Hubei (0.1) | 96 (10-113) | 0.100 (0.100-0.100) | 5.148 (1.690-9.625) | 0.034 (0.033-0.046) |
| Hubei (0.2) | 42 (41-52) | 0.200 (0.200-0.200) | 8.593 (5.221-9.625) | 0.026 (0.023-0.026) |
| Hubei (0.3) | 23 (7-24) | 0.300 (0.300-0.343) | 5.635 (0.510-12.356) | 0.018 (0.017-0.028) |
| | RMSE | $R_0$ | DT | $R'_0$ |
| Hubei | 405.544 (389.821-448.441) | 3.850 (3.083-4.595) | 2.625 (2.217-3.267) | 0.499 (0.489-0.520) |
| Hubei (0.1) | 558.091 (490.407-564.010) | 2.204 (2.164-3.040) | 4.069 (2.688-4.174) | 0.586 (0.556-0.590) |
| Hubei (0.2) | 673.229 (670.861-687.861) | 1.438 (1.392-1.444) | 7.217 (7.124-7.937) | 0.623 (0.621-0.626) |
| Hubei (0.3) | 750.531 (681.687-758.662) | 1.010 (0.996-1.143) | 134.995 (7.051-408.040) | 0.597 (0.565-0.599) |

## B Details of the SEIR model

The SEIR model consists of four compartments: *S* for the number of susceptible, *E* for the number of exposed, *I* for the number of infectious, and *R* for the number recovered (or immune) individuals. The dynamics is described by the following differential equations:

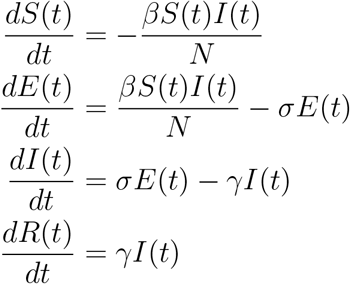

where *β* represents the rate of transmission for the susceptible to be infected; *σ* is the rate by which the exposed individual develops symptoms; *γ* is the probability of recovery or death; and *N* is the total population.

In order to apply the SEIR model, we need to estimate the parameters *β, σ*, and *γ*. Because the incubation period of the COVID-19 has been reported to be between 2 to 14 days, we chose the midpoint of 7 days, and *σ* is set to be 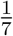. *γ* is the average rate of recovery or death in infected populations (i.e., 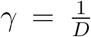, where *D* is the average duration of the infection). According to the published clinical experience, we set *D* to be 14 and 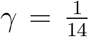. The parameter *β* and the number of initial infections can be learned to fit the daily COVID-19 outbreak data. We assume that the value of *β* is reduced by 80% after Jan. 23 because of the containment. We assume the diagnostic period is 10 days, which means the number of new infections corresponds to the number of newly confirmed cases 10 days later.

## C LSTM Training Details

The LSTM model implemented in this paper takes the number of newly confirmed cases in the previous three days as input, and output the number of newly confirmed cases in the next day. The architecture of the neural network is: Input(3) → FC(3,15) → Tanh → LSTM(15,15) → FC(15,1) → →Tanh →output(1), where FC means a fully connected layer, tanh means tanh activation function and LSTM means an LSTM module.

An example was made by taking the previous three days’ number of confirmed cases as the feature and this day’s number as the label. All examples consisted the training set. The values in a feature were rescaled to [0, 1] by min-max normalization. In each training iteration, an episode of features was inputted, and the mean square error between the outputs and the labels were calculated. We used Adam to optimize the neural network parameters. The learning rate was set to be 0.001, and the neural network was optimized for 500 iterations. In the test phase, the current output of the model was used as the future input for the lack of real data.

## D Parameter settings for the optimization

Here, we list the parameter settings for the optimization in all experiments. We set the range of the parameter’s search space to a reasonable range according to the prior knowledge. *γ*, the diagnostic rate per day, was upper bounded by 0.3. Considering the overall mortality rate of COVID-19 was no more than 20%, we upper-bounded *δ*, the mortality rate per day, by 1%. *k*, the average contacts per person per day after the containment, was bounded by 15, which is the average contacts before the containment. *θ*, the recovery rate per day of the confirmed cases, was bounded by 10% for China or 15% for other countries. *I*(0), the number of initial infections 14 days before the date when the epidemic data was first released, was bounded by 400 for Hubei, China, and other countries. *I*(0) was set to be 0 for other provinces in China because the initial infections in these provinces were from Hubei. If *β* is 0.1, the value of *R*_0_ will be 9.16, which is far from the *R*_0_ value determined by current literature. Thus, *β* of Hubei was upper bounded by 0.1. Budget, the number of evaluations the optimization algorithm can use, was set to be 200,000 for the optimization of parameters in Hubei. For other provinces and countries, the budget was set to be 80,000 to reduce the total training time.

For the remaining parameters, we first set their search space to a large and reasonable range. The range of *σ*, the quarantine rate per day, is set to be [0, 0.24]. The average contacts *k* after the containment was upper bounded by 15. We first optimized the parameters of Hubei. Then we further narrow down the search space according to the obtained parameters. For example, *β* was limited to [0, 0.5], and *k* was limited to [0, 5]. The ultimate search space of all parameters was shown in Table 2.

**Table 2:** Parameter settings for the optimization.

| Experiment | Options | $\beta$ | $k$ | $\gamma$ | $\delta$ | $I(0)$ | $\sigma$ | $k'$ | $\theta$ | budget |
| --- | --- | --- | --- | --- | --- | --- | --- | --- | --- | --- |
| Comparision | Hubei | $[0, 0.1]$ | $[0, 5]$ | $[0.0, 0.3]$ | $[0.0, 0.01]$ | $[0, 400]$ | $[0.03, 0.12]$ | $[0, 15]$ | $[0.0, 0.1]$ | 200000 |
| | Exclude Hubei | $[0, 0.05]$ | $[0, 5]$ | $[0.0, 0.3]$ | $[0.0, 0.01]$ | 0 | $[0.03, 0.12]$ | $[0, 15]$ | $[0.0, 0.1]$ | 80000 |
| | South Korea | $[0, 0.05]$ | $[0, 5]$ | $[0.0, 0.3]$ | $[0.0, 0.01]$ | $[0, 400]$ | $[0.03, 0.12]$ | $[0, 15]$ | $[0.0, 0.15]$ | 80000 |
| Forecast China's mainland | Hubei | $[0, 0.1]$ | $[0, 5]$ | $[0.0, 0.3]$ | $[0.0, 0.01]$ | $[0, 400]$ | $[0.03, 0.24]$ | $[0, 15]$ | $[0.0, 0.1]$ | 200000 |
| | Exclude Hubei | $[0, 0.05]$ | $[0, 5]$ | $[0.0, 0.3]$ | $[0.0, 0.01]$ | 0 | $[0.03, 0.12]$ | $[0, 15]$ | $[0.0, 0.1]$ | 80000 |
| Forecast Other countries | Obtain $k$ | $[0, 0.05]$ | $[0, 5]$ | $[0.0, 0.3]$ | $[0.0, 0.01]$ | $[0, 400]$ | $[0.03, 0.12]$ | $[0, 15]$ | $[0.0, 0.15]$ | 80000 |
| | Fix $k$ | $[0, 0.05]$ | 15 | $[0.0, 0.3]$ | $[0.0, 0.01]$ | $[0, 400]$ | $[0.03, 0.12]$ | $[0, 15]$ | $[0.0, 0.15]$ | 80000 |

## E The simulated results of the number of infections on April 8

**Table 3:** The simulated results of the number of all kinds of infections on April 15. The number of infections in China’s mainland is the sum of infections in 31 provinces. The optimization was repeated for 10 times and the median and its confidence interval was recorded.

|  | Real confirmed | MLSim confirmed | Total infections | Latent | Self-healed |
| --- | --- | --- | --- | --- | --- |
| China's mainland | 82,249 | 84,314 (82,485-86,920) | 233,047 (221,551-248,305) | 121 (94-174) | 135,993 (126,287-146,838) |
| Beijing | 589 | 400 (394-417) | 596 (545-670) | 0 (0-0) | 104 (78-185) |
| Tianjin | 185 | 133 (128-152) | 198 (184-267) | 0 (0-0) | 42 (27-91) |
| Hebei | 327 | 359 (351-382) | 702 (556-1,032) | 0 (0-1) | 215 (137-480) |
| Shanxi | 173 | 135 (129-146) | 208 (182-223) | 0 (0-0) | 45 (29-67) |
| InnerMongoria | 190 | 81 (78-89) | 118 (109-137) | 0 (0-0) | 26 (19-47) |
| Liaoning | 145 | 134 (127-146) | 285 (232-339) | 0 (0-0) | 80 (45-112) |
| Jilin | 100 | 98 (93-104) | 123 (117-163) | 0 (0-0) | 22 (20-53) |
| Heilongjiang | 819 | 513 (500-524) | 713 (643-894) | 0 (0-0) | 182 (102-318) |
| Shanghai | 618 | 356 (347-375) | 546 (477-808) | 0 (0-0) | 147 (90-372) |
| Jiangsu | 653 | 704 (678-722) | 1,326 (1,045-1,810) | 0 (0-1) | 450 (200-836) |
| Zhejiang | 1,267 | 1,256 (1,233-1,288) | 2,033 (1,604-2,547) | 0 (0-0) | 702 (251-1,152) |
| Anhui | 991 | 1,082 (1,055-1,114) | 1,621 (1,518-2,002) | 0 (0-0) | 401 (335-729) |
| Fujian | 353 | 323 (302-327) | 472 (411-576) | 0 (0-0) | 141 (66-219) |
| Jiangxi | 937 | 1,070 (1,040-1,086) | 1,462 (1,342-2,346) | 0 (0-0) | 319 (213-1,191) |
| Shandong | 784 | 730 (716-750) | 1,150 (1,076-1,264) | 1 (1-1) | 296 (219-373) |
| Henan | 1,276 | 1,483 (1,465-1,515) | 2,911 (2,275-3,446) | 1 (1-1) | 1,249 (623-1,753) |
| Hubei | 67,803 | 70,316 (68,858-72,492) | 208,793 (200,417-218,028) | 116 (90-165) | 127,815 (120,864-133,428) |
| Hunan | 1,019 | 1,235 (1,216-1,261) | 3,655 (3,521-3,745) | 2 (2-3) | 2,168 (2,061-2,226) |
| Guangdong | 1,564 | 1,384 (1,348-1,409) | 1,924 (1,772-2,276) | 0 (0-0) | 378 (253-705) |
| Guangxi | 254 | 264 (246-270) | 354 (335-412) | 0 (0-0) | 85 (61-142) |
| Hainan | 168 | 177 (157-186) | 292 (247-457) | 0 (0-0) | 87 (55-254) |
| Chongqing | 579 | 649 (635-661) | 1,087 (947-1,663) | 0 (0-1) | 334 (212-821) |
| Sichuan | 560 | 573 (566-594) | 943 (784-1,167) | 0 (0-1) | 232 (111-415) |
| Guizhou | 146 | 151 (148-155) | 245 (205-311) | 0 (0-0) | 70 (40-134) |
| Yunnan | 184 | 175 (168-194) | 326 (238-453) | 0 (0-0) | 80 (26-165) |
| Tibet | 1 | 8 (7-8) | 10 (9-14) | 0 (0-0) | 2 (2-6) |
| Shanxi | 256 | 261 (257-274) | 532 (435-691) | 0 (0-0) | 177 (94-291) |
| Gansu | 139 | 90 (86-99) | 145 (123-185) | 0 (0-0) | 48 (20-88) |
| Qinghai | 18 | 15 (15-16) | 23 (20-27) | 0 (0-0) | 6 (4-10) |
| Ningxia | 75 | 76 (68-85) | 125 (89-193) | 0 (0-0) | 41 (15-94) |
| Xinjiang | 76 | 76 (74-79) | 123 (93-159) | 0 (0-0) | 42 (15-81) |
| SouthKorea | 10,564 | 10,443 (9,950-11,508) | 20,708 (13,529-31,790) | 112 (40-262) | 10,232 (3,356-20,455) |
| USA | 607,670 | 658,177 (650,044-663,172) | 2,136,228 (1,807,158-2,622,230) | 354,657 (277,641-495,128) | 977,795 (637,699-1,311,312) |
| Italy | 162,488 | 150,623 (150,167-151,686) | 501,153 (389,010-522,253) | 41,387 (29,037-57,151) | 294,992 (154,536-297,085) |
| UK | 93,873 | 103,068 (99,525-107,401) | 320,425 (304,455-394,350) | 144,424 (127,215-171,930) | 48,518 (45,303-59,118) |
| France | 130,253 | 371,852 (266,108-533,929) | 1,259,440 (408,079-1,668,240) | 40,379 (10,807-186,878) | 679,208 (81,067-802,794) |
| Germany | 131,359 | 145,306 (140,573-151,058) | 253,951 (208,341-419,828) | 21,118 (11,484-41,646) | 87,570 (44,031-226,567) |

## F Obtained simulator parameters by fitting the COVID-10 data

**Table 4:** Obtained parameters by fitting the COVID-19 data. For Chinese provinces, data before March 13 was used for training and for other countries, data before April 8 was used. Optimization were repeated for 10 times and the median and its confidence interval was recorded. *β* is the infection rate. *k* is the average number of contacts after containment. The average number of contacts is 15 before containment. *γ* is diagnostic rate per day. *δ* is the mortality rate per day.

| | $\beta$ | $k$ | $\gamma$ | $\delta$ |
| --- | --- | --- | --- | --- |
| Beijing | 0.031 (0.026-0.036) | 1.728 (0.778-2.289) | 0.129 (0.096-0.172) | 0.005 (0.005-0.006) |
| Tianjin | 0.042 (0.035-0.049) | 1.070 (0.551-1.913) | 0.065 (0.048-0.110) | 0.003 (0.000-0.004) |
| Hebei | 0.034 (0.025-0.039) | 0.285 (0.119-0.769) | 0.108 (0.084-0.132) | 0.001 (0.001-0.002) |
| Shanxi | 0.029 (0.023-0.034) | 0.757 (0.239-2.020) | 0.098 (0.077-0.128) | 0.004 (0.002-0.005) |
| InnerMongoria | 0.042 (0.035-0.049) | 1.205 (0.723-2.148) | 0.120 (0.109-0.145) | 0.002 (0.000-0.004) |
| Liaoning | 0.030 (0.030-0.030) | 0.023 (0.000-0.144) | 0.299 (0.298-0.300) | 0.004 (0.000-0.007) |
| Jilin | 0.048 (0.038-0.049) | 0.933 (0.122-1.452) | 0.039 (0.029-0.054) | 0.004 (0.001-0.006) |
| Heilongjiang | 0.048 (0.043-0.050) | 1.249 (0.918-1.435) | 0.026 (0.020-0.035) | 0.003 (0.002-0.004) |
| Shanghai | 0.035 (0.028-0.041) | 0.333 (0.134-0.616) | 0.152 (0.116-0.248) | 0.002 (0.001-0.002) |
| Jiangsu | 0.030 (0.024-0.036) | 0.268 (0.090-0.431) | 0.087 (0.080-0.112) | 0.001 (0.001-0.003) |
| Zhejiang | 0.031 (0.029-0.037) | 0.704 (0.251-1.189) | 0.154 (0.124-0.158) | 0.004 (0.003-0.005) |
| Anhui | 0.023 (0.020-0.030) | 0.386 (0.309-0.566) | 0.051 (0.045-0.075) | 0.000 (0.000-0.001) |
| Fujian | 0.025 (0.019-0.029) | 0.335 (0.051-0.886) | 0.159 (0.132-0.190) | 0.001 (0.000-0.001) |
| Jiangxi | 0.019 (0.011-0.028) | 0.051 (0.024-0.154) | 0.063 (0.038-0.082) | 0.002 (0.001-0.002) |
| Shandong | 0.032 (0.027-0.038) | 1.439 (1.090-2.069) | 0.163 (0.126-0.188) | 0.001 (0.000-0.001) |
| Henan | 0.016 (0.014-0.019) | 0.051 (0.005-0.134) | 0.145 (0.111-0.164) | 0.002 (0.001-0.002) |
| Hubei | 0.023 (0.018-0.027) | 1.950 (1.572-2.533) | 0.000 (0.000-0.000) | 0.005 (0.005-0.005) |
| Hunan | 0.006 (0.001-0.011) | 0.025 (0.006-0.149) | 0.300 (0.299-0.300) | 0.005 (0.004-0.006) |
| Guangdong | 0.020 (0.017-0.029) | 0.795 (0.652-1.598) | 0.121 (0.090-0.140) | 0.004 (0.004-0.005) |
| Guangxi | 0.029 (0.020-0.033) | 1.644 (0.840-2.347) | 0.125 (0.108-0.170) | 0.002 (0.001-0.002) |
| Hainan | 0.040 (0.034-0.048) | 0.842 (0.096-1.288) | 0.119 (0.090-0.151) | 0.002 (0.001-0.003) |
| Chongqing | 0.007 (0.003-0.013) | 0.414 (0.089-1.328) | 0.269 (0.203-0.289) | 0.003 (0.002-0.004) |
| Sichuan | 0.012 (0.007-0.015) | 1.336 (0.497-2.321) | 0.135 (0.110-0.165) | 0.003 (0.001-0.004) |
| Guizhou | 0.022 (0.016-0.028) | 0.516 (0.311-1.026) | 0.038 (0.018-0.048) | 0.004 (0.003-0.004) |
| Yunnan | 0.031 (0.029-0.033) | 0.910 (0.128-1.747) | 0.227 (0.168-0.271) | 0.001 (0.001-0.003) |
| Tibet | 0.034 (0.031-0.037) | 0.465 (0.242-1.349) | 0.096 (0.007-0.132) | 0.004 (0.002-0.008) |
| Shanxi | 0.014 (0.008-0.022) | 0.304 (0.142-0.671) | 0.140 (0.121-0.161) | 0.002 (0.000-0.003) |
| Gansu | 0.028 (0.023-0.032) | 0.150 (0.070-0.821) | 0.167 (0.116-0.204) | 0.004 (0.003-0.007) |
| Qinghai | 0.031 (0.026-0.038) | 0.261 (0.037-1.027) | 0.247 (0.233-0.296) | 0.007 (0.004-0.009) |
| Ningxia | 0.043 (0.034-0.049) | 1.227 (0.470-1.990) | 0.148 (0.110-0.178) | 0.007 (0.001-0.009) |
| Xinjiang | 0.040 (0.036-0.045) | 1.026 (0.564-1.445) | 0.062 (0.031-0.072) | 0.002 (0.000-0.004) |
| SouthKorea | 0.015 (0.011-0.022) | 3.228 (2.721-4.733) | 0.299 (0.256-0.300) | 0.005 (0.004-0.006) |
| USA | 0.011 (0.011-0.012) | 5.000 (4.971-5.000) | 0.027 (0.008-0.065) | 0.005 (0.004-0.005) |
| Italy | 0.012 (0.012-0.016) | 5.000 (5.000-5.000) | 0.000 (0.000-0.012) | 0.010 (0.010-0.010) |
| UK | 0.011 (0.010-0.011) | 14.985 (14.985-14.986) | 0.214 (0.058-0.283) | 0.010 (0.010-0.010) |
| France | 0.016 (0.014-0.020) | 2.335 (0.559-2.871) | 0.016 (0.001-0.088) | 0.004 (0.004-0.005) |
| Germany | 0.012 (0.010-0.014) | 4.385 (4.013-5.000) | 0.299 (0.192-0.300) | 0.003 (0.003-0.003) |

**Table 5:** Obtained parameters by fitting the COVID-19 data. Follow Table 4. For Chinese provinces, *I*(0) is the number of initial infections on Dec. 28, 2019. For other countries, *I*(0) is the number of initial infections on Jan. 9, 2020. *σ* is the quarantine rate per day. *k* is the average number of contacts in the cross-region population movement. *γ* is diagnostic rate per day. *θ* is the recovery rate per day.

| | $I(0)$ | $\sigma$ | $k'$ | $\theta$ |
| --- | --- | --- | --- | --- |
| Beijing | 0 (0-0) | 0.102 (0.074-0.115) | 12.652 (5.712-14.701) | 0.079 (0.070-0.088) |
| Tianjin | 0 (0-0) | 0.095 (0.065-0.114) | 2.164 (0.124-10.539) | 0.099 (0.090-0.100) |
| Hebei | 0 (0-0) | 0.061 (0.034-0.076) | 2.223 (0.405-8.912) | 0.100 (0.100-0.100) |
| Shanxi | 0 (0-0) | 0.090 (0.068-0.101) | 7.359 (4.039-13.992) | 0.089 (0.082-0.100) |
| InnerMongoria | 0 (0-0) | 0.093 (0.064-0.109) | 6.107 (1.617-12.561) | 0.002 (0.000-0.013) |
| Liaoning | 0 (0-0) | 0.054 (0.046-0.077) | 1.953 (0.038-4.112) | 0.098 (0.096-0.100) |
| Jilin | 0 (0-0) | 0.112 (0.073-0.120) | 11.678 (5.002-14.971) | 0.088 (0.063-0.099) |
| Heilongjiang | 0 (0-0) | 0.088 (0.066-0.115) | 132.513 (66.229-144.317) | 0.099 (0.091-0.100) |
| Shanghai | 0 (0-0) | 0.080 (0.045-0.103) | 5.794 (1.594-9.703) | 0.099 (0.095-0.100) |
| Jiangsu | 0 (0-0) | 0.057 (0.037-0.094) | 10.358 (8.274-13.620) | 0.100 (0.100-0.100) |
| Zhejiang | 0 (0-0) | 0.068 (0.050-0.119) | 9.806 (5.861-13.524) | 0.077 (0.074-0.083) |
| Anhui | 0 (0-0) | 0.087 (0.062-0.094) | 10.221 (5.699-14.317) | 0.099 (0.098-0.100) |
| Fujian | 0 (0-0) | 0.077 (0.060-0.113) | 4.522 (0.418-8.211) | 0.045 (0.041-0.055) |
| Jiangxi | 0 (0-0) | 0.101 (0.042-0.118) | 9.639 (4.971-14.514) | 0.100 (0.100-0.100) |
| Shandong | 0 (0-0) | 0.077 (0.069-0.094) | 9.343 (4.101-13.511) | 0.051 (0.045-0.057) |
| Henan | 0 (0-0) | 0.053 (0.041-0.081) | 1.759 (0.599-7.308) | 0.100 (0.100-0.100) |
| Hubei | 106 (29-397) | 0.030 (0.030-0.031) | 4.246 (1.430-12.966) | 0.049 (0.042-0.054) |
| Hunan | 0 (0-0) | 0.031 (0.030-0.032) | 0.359 (0.049-14.544) | 0.100 (0.100-0.100) |
| Guangdong | 0 (0-0) | 0.100 (0.070-0.119) | 6.689 (3.523-11.827) | 0.093 (0.091-0.099) |
| Guangxi | 0 (0-0) | 0.095 (0.067-0.110) | 5.966 (0.774-13.292) | 0.043 (0.032-0.051) |
| Hainan | 0 (0-0) | 0.076 (0.036-0.094) | 7.558 (3.220-14.396) | 0.096 (0.088-0.099) |
| Chongqing | 0 (0-0) | 0.070 (0.039-0.091) | 8.459 (1.168-12.931) | 0.097 (0.094-0.100) |
| Sichuan | 0 (0-0) | 0.077 (0.053-0.112) | 8.040 (3.396-12.182) | 0.064 (0.055-0.072) |
| Guizhou | 0 (0-0) | 0.080 (0.049-0.101) | 12.469 (1.861-13.923) | 0.099 (0.097-0.100) |
| Yunnan | 0 (0-0) | 0.068 (0.039-0.115) | 6.090 (1.678-10.838) | 0.037 (0.027-0.040) |
| Tibet | 0 (0-0) | 0.101 (0.055-0.116) | 4.143 (1.403-10.461) | 0.035 (0.013-0.086) |
| Shanxi | 0 (0-0) | 0.054 (0.038-0.082) | 6.702 (3.025-12.939) | 0.086 (0.075-0.092) |
| Gansu | 0 (0-0) | 0.070 (0.047-0.119) | 4.849 (0.140-12.051) | 0.099 (0.098-0.100) |
| Qinghai | 0 (0-0) | 0.083 (0.061-0.107) | 9.237 (0.246-14.057) | 0.099 (0.093-0.100) |
| Ningxia | 0 (0-0) | 0.070 (0.043-0.113) | 6.160 (0.994-12.243) | 0.095 (0.081-0.100) |
| Xinjiang | 0 (0-0) | 0.070 (0.044-0.119) | 7.138 (2.773-13.084) | 0.025 (0.011-0.043) |
| SouthKorea | 5 (0-80) | 0.052 (0.031-0.094) | 3.911 (0.615-12.778) | 0.016 (0.002-0.023) |
| USA | 205 (72-235) | 0.038 (0.030-0.048) | 7.670 (0.065-13.656) | 0.000 (0.000-0.000) |
| Italy | 68 (6-122) | 0.030 (0.030-0.049) | 7.410 (0.310-11.794) | 0.000 (0.000-0.003) |
| UK | 94 (59-130) | 0.067 (0.067-0.068) | 7.190 (2.711-8.759) | 0.000 (0.000-0.000) |
| France | 1 (0-4) | 0.030 (0.030-0.100) | 4.204 (0.834-12.302) | 0.000 (0.000-0.000) |
| Germany | 53 (11-132) | 0.064 (0.034-0.099) | 7.773 (2.450-10.517) | 0.150 (0.150-0.150) |

**Table 6:** Obtained parameters by fitting the COVID-19 data. Follow Table 5. RMSE is the root mean square error between the real data and the simulated results. *R*_0_ and 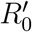 is the reproductive number before and after the containment. *DT* is the epidemic doubling time in the early stage, which ignores the self-healed patients.

| | RMSE | $R_0$ | DT | $R'_0$ |
| --- | --- | --- | --- | --- |
| Beijing | 3.479 (3.416-3.575) | 3.147 (2.698-4.146) | 2.543 (2.119-3.036) | 0.390 (0.237-0.417) |
| Tianjin | 1.837 (1.777-1.923) | 4.507 (3.339-6.105) | 1.789 (1.465-2.344) | 0.317 (0.180-0.490) |
| Hebei | 3.853 (3.720-3.954) | 4.886 (3.185-5.869) | 1.929 (1.753-2.773) | 0.105 (0.027-0.261) |
| Shanxi | 1.244 (1.222-1.369) | 3.557 (2.531-4.128) | 2.409 (2.049-3.381) | 0.211 (0.060-0.353) |
| InnerMongoria | 1.098 (1.069-1.155) | 4.864 (3.404-6.170) | 1.732 (1.486-2.283) | 0.353 (0.287-0.473) |
| Liaoning | 2.489 (2.211-2.679) | 4.249 (3.631-4.493) | 2.198 (2.139-2.382) | 0.006 (0.000-0.043) |
| Jilin | 1.447 (1.420-1.507) | 4.521 (3.619-5.220) | 1.645 (1.590-2.089) | 0.263 (0.061-0.336) |
| Heilongjiang | 5.083 (5.014-5.258) | 5.066 (4.550-6.059) | 1.610 (1.446-1.699) | 0.399 (0.371-0.465) |
| Shanghai | 3.814 (3.740-3.958) | 4.004 (3.081-5.954) | 2.112 (1.596-2.803) | 0.101 (0.030-0.236) |
| Jiangsu | 4.108 (3.934-4.665) | 4.377 (2.563-5.741) | 2.179 (1.767-3.383) | 0.077 (0.020-0.156) |
| Zhejiang | 15.787 (15.463-15.904) | 4.202 (2.789-5.265) | 2.199 (1.772-2.792) | 0.187 (0.075-0.221) |
| Anhui | 7.281 (7.077-7.394) | 2.693 (2.088-3.870) | 3.273 (2.315-4.533) | 0.075 (0.057-0.104) |
| Fujian | 2.810 (2.746-2.867) | 2.824 (2.030-4.196) | 3.176 (2.195-4.585) | 0.055 (0.015-0.130) |
| Jiangxi | 7.749 (7.525-8.022) | 2.064 (1.042-4.351) | 4.486 (2.238-23.097) | 0.008 (0.002-0.029) |
| Shandong | 5.965 (5.598-6.461) | 3.763 (3.028-4.387) | 2.338 (1.923-2.716) | 0.370 (0.265-0.411) |
| Henan | 9.901 (9.238-10.228) | 2.271 (1.949-2.664) | 4.017 (2.599-5.068) | 0.006 (0.001-0.017) |
| Hubei | 405.544 (389.821-448.441) | 3.850 (3.083-4.595) | 2.625 (2.217-3.267) | 0.499 (0.489-0.520) |
| Hunan | 8.955 (8.720-9.437) | 1.035 (0.086-1.857) | 7.608 (-24.491-32.180) | 0.001 (0.000-0.004) |
| Guangdong | 8.288 (7.791-8.811) | 1.978 (1.737-3.702) | 4.686 (2.391-5.832) | 0.173 (0.077-0.213) |
| Guangxi | 2.823 (2.795-2.845) | 3.207 (1.984-4.757) | 2.536 (1.992-4.635) | 0.346 (0.199-0.423) |
| Hainan | 2.191 (2.142-2.225) | 5.250 (4.017-6.706) | 1.753 (1.471-2.093) | 0.323 (0.041-0.365) |
| Chongqing | 4.896 (4.816-5.000) | 0.878 (0.324-1.986) | -1.501 (-31.704-8.640) | 0.020 (0.001-0.035) |
| Sichuan | 3.647 (3.588-3.742) | 1.266 (0.628-2.083) | 5.364 (-19.439-14.843) | 0.135 (0.039-0.203) |
| Guizhou | 2.365 (2.303-2.407) | 2.687 (1.644-4.359) | 3.461 (2.236-6.452) | 0.110 (0.054-0.139) |
| Yunnan | 2.486 (2.462-2.548) | 3.845 (3.017-4.682) | 2.320 (1.894-2.654) | 0.203 (0.034-0.355) |
| Tibet | 0.000 (0.000-0.000) | 3.628 (2.878-4.609) | 2.161 (2.027-2.721) | 0.123 (0.066-0.330) |
| Shanxi | 2.446 (2.350-2.488) | 1.880 (1.095-3.553) | 5.415 (2.765-13.693) | 0.042 (0.017-0.066) |
| Gansu | 3.117 (3.065-3.135) | 3.509 (2.417-4.562) | 2.627 (2.097-3.386) | 0.038 (0.018-0.159) |
| Qinghai | 0.471 (0.471-0.471) | 3.347 (2.776-4.845) | 2.489 (1.830-2.996) | 0.056 (0.013-0.228) |
| Ningxia | 1.225 (1.208-1.247) | 5.430 (4.044-6.317) | 1.638 (1.543-2.187) | 0.419 (0.165-0.529) |
| Xinjiang | 1.257 (1.168-1.285) | 5.316 (3.713-6.220) | 1.745 (1.606-2.011) | 0.356 (0.234-0.408) |
| SouthKorea | 98.208 (93.090-117.351) | 2.129 (1.655-2.393) | 4.539 (3.862-6.473) | 0.482 (0.439-0.496) |
| USA | 1933.981 (1626.672-2128.964) | 1.798 (1.699-2.000) | 5.876 (5.188-6.314) | 0.599 (0.566-0.660) |
| Italy | 568.740 (435.673-678.736) | 2.046 (1.961-2.379) | 5.003 (4.285-5.308) | 0.682 (0.654-0.793) |
| UK | 698.713 (695.328-709.456) | 1.385 (1.344-1.417) | 8.764 (8.427-9.241) | 1.383 (1.342-1.416) |
| France | 6516.358 (3754.157-10637.327) | 2.420 (2.012-2.684) | 4.207 (3.660-4.651) | 0.323 (0.091-0.459) |
| Germany | 669.210 (631.233-728.894) | 1.583 (1.349-1.675) | 6.949 (6.398-9.585) | 0.456 (0.445-0.489) |

## G Simulation of the virus transmission in Chinese provinces with the obtained parameters

**Figure 6:**
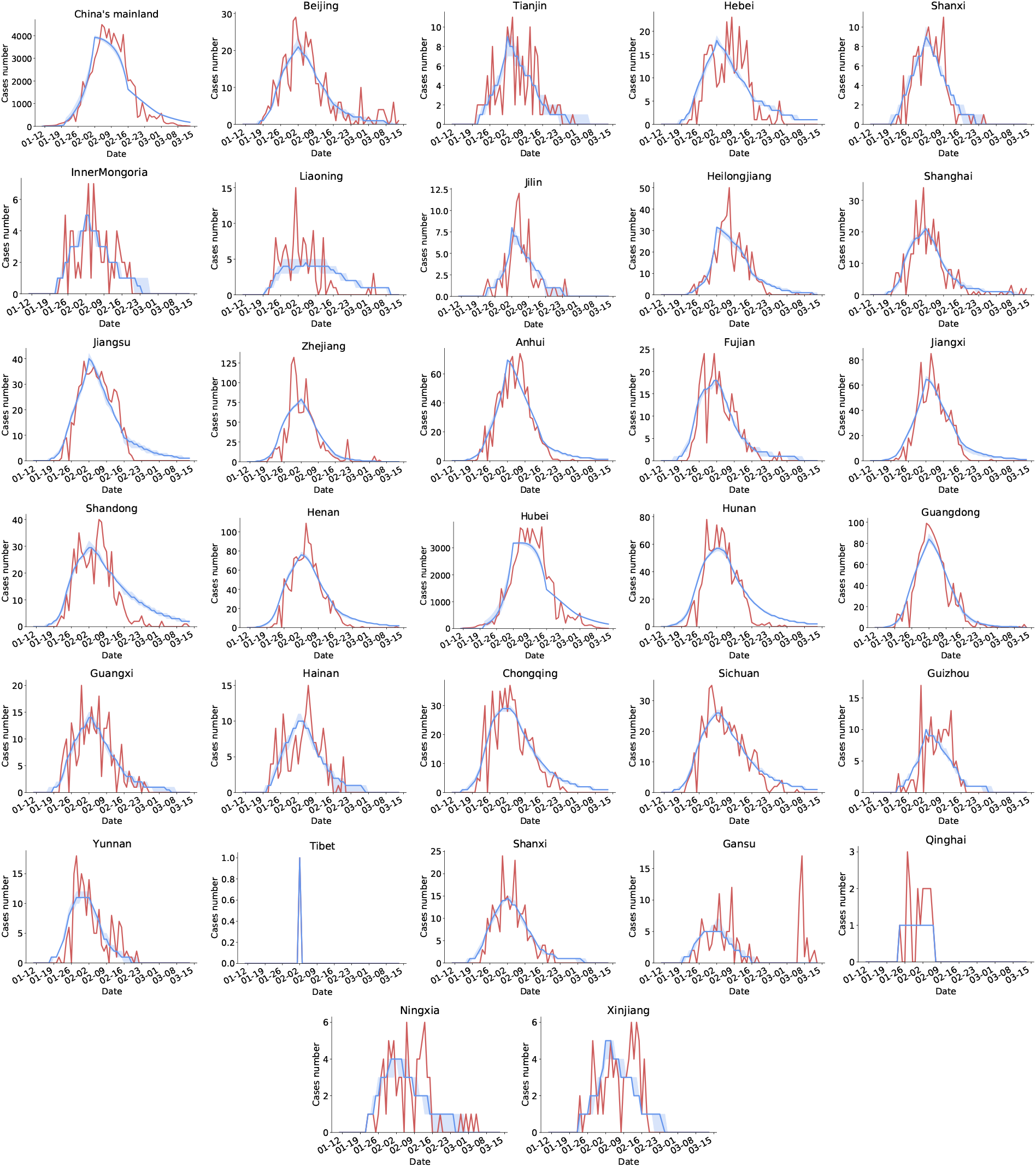
The comparison between the real (red) number of newly confirmed cases and the simulated (blue) results. It can be observed that MLSim can fit the real data well.

## Footnotes

1 https://github.com/BlankerL/DXY-COVID-19-Data

2 https://github.com/CSSEGISandData/COVID-19

3 http://qianxi.baidu.com

